# Access to Sexual and Reproductive Health Education and Services Among Deaf Adolescents in Wakiso District, Uganda: A Mixed-Methods Cross-Sectional Study

**DOI:** 10.64898/2026.05.27.26354296

**Authors:** Racheal Ayanga, Nancy Katumba Muwangala, Jane Babirye, Robert Nkwangu

## Abstract

**Background:** Persons with disabilities, particularly deaf individuals, remain a largely overlooked population in sexual and reproductive health (SRH) programming globally, with this gap especially pronounced in low- and middle-income countries. Deafness imposes substantial barriers to accessing information and services that are routinely available to hearing peers, further exacerbated in the post-COVID-19 era. This study assessed deaf adolescents’ knowledge of and access to SRH education and services in Wakiso District, Uganda, and explored systemic, institutional, community, and adolescent-level factors shaping access.

**Methods:** A mixed-methods cross-sectional study was conducted at Wakiso Secondary School for the Deaf from July 2022 to January 2023. Quantitative data were collected from 70 consecutively sampled deaf adolescents aged 13–19 years using a structured questionnaire. Qualitative data were gathered through key informant interviews (KIIs) with four purposively selected stakeholders and a focus group discussion (FGD) with deaf adolescent students. Qualitative data were analysed thematically.

**Results:** The mean participant age was 17 years (SD ±1.8); 65.7% were female. A large majority (88.6%) had heard of SRH components, and 98.6% perceived a need for SRH education or services. However, 84.3% reported challenges accessing these services at least 85% of the time. No participant had ever received SRH education or services through a formal health facility. The FGD revealed that adolescents’ conceptualisation of SRH was narrow, centred on body hygiene and HIV prevention, while service-seeking was reactive and symptom-driven. Five cross-cutting themes emerged from the KIIs and were reinforced by FGD findings: communication barriers; inadequate and inaccessible services; family and community isolation; existing platforms and positive practices; and negative provider attitudes and limited capacity. The school nurse emerged as the sole functional SRH access point for most participants.

**Conclusion:** Despite high awareness and near-universal perceived need, deaf adolescents in Uganda face profound multilevel barriers to SRH access. Structural, psychosocial, and knowledge-related barriers interact to exclude this population from formal health services. Findings call for disability-responsive SRH integration into health systems, training of health workers in accessible communication, community capacity building, and co-design of SRH programmes with deaf adolescents.

## 1. Background and Introduction

Disability affects more than one billion people globally, approximately 15.6% of the world’s population and is disproportionately prevalent among women, people in poverty, and residents of low- and middle-income countries (LMICs) [1]. Poverty both causes and is perpetuated by disability through overlapping barriers in communication, employment, education, and access to health care. Despite the commitment of the United Nations’ Sustainable Development Goals (SDGs) to leave no one behind, persons with disabilities remain systematically excluded from health programming, including sexual and reproductive health (SRH) [2].

Among the diverse spectrum of disability, deafness presents a unique and frequently underappreciated set of challenges in the SRH domain. Deaf individuals rely on visual and manual modes of communication, primarily sign language, that are poorly understood or unavailable within most health systems. For adolescents, the transition through puberty and early sexual development already presents biological, psychological, and social complexity. Deaf adolescents must navigate this transition without access to the ambient health information available to hearing peers through radio, conversation, and formal classroom instruction, and with the compounding effects of communication exclusion within their families and communities.

The COVID-19 pandemic widened these disparities substantially. Movement restrictions limited community services; mask-wearing obscured lip movements critical to deaf communication; and the rapid pivot to digital health information assumed literacy and internet access rarely available to deaf adolescents in peri-urban Uganda [3]. These conditions rendered an already underserved population even more isolated from SRH information and care.

Uganda, a lower-middle-income country in sub-Saharan Africa, has one of the youngest populations in the world, with approximately 78% of its citizens below 30 years of age [4]. Adolescent SRH remains a national public health priority, given high rates of teenage pregnancy, sexually transmitted infections (STIs), and limited uptake of contraception [5]. However, disability-inclusive SRH programming is largely absent from Uganda’s national health policy and service delivery landscape. Deaf adolescents are rarely included as target groups in national SRH education campaigns, and health workers are seldom trained in sign language or deaf-accessible communication.

Despite these structural realities, there is a paucity of empirical data documenting the SRH knowledge, experiences, and access barriers of deaf adolescents in Uganda. Most existing evidence on disability and SRH is drawn from high-income contexts and does not reflect the specific socio-cultural, linguistic, and systems-level barriers present in sub-Saharan Africa. Critically, the voices of deaf adolescents themselves are often absent from research, limiting the depth and authenticity of findings. This evidence gap constrains the ability of policymakers and service providers to develop targeted, effective, and equitable responses.

This study was designed to address this gap using a mixed-methods approach. Quantitative data capture the prevalence and nature of SRH knowledge and access barriers among deaf adolescents; qualitative key informant interviews provide institutional and community stakeholder perspectives; and a focus group discussion (FGD) centres the voices and lived experiences of deaf adolescents themselves. Together, the three data strands offer a comprehensive, multi-perspective picture of the problem and a stronger evidence base for action.

## 2. Methods

### 2.1 Study Design

We conducted a convergent mixed-methods cross-sectional study, integrating quantitative survey data with qualitative data from key informant interviews (KIIs) and a focus group discussion (FGD) with deaf adolescents. The three components were conducted concurrently during the same study period, with integration occurring at the interpretation stage. The mixed-methods design was chosen to triangulate findings, explore mechanisms underlying quantitative patterns, and capture both the lived institutional realities of SRH provision and the first-person experiential perspectives of deaf adolescents.

### 2.2 Study Setting and Period

The study was conducted at Wakiso Secondary School for the Deaf (WSSD), located in Wakiso District, central Uganda, from 01 July 2022 to 30 June 2023. WSSD is one of Uganda’s few government-supported schools specifically serving deaf students, providing a concentrated and accessible sample of deaf adolescents and key institutional actors. Wakiso District borders Kampala, the national capital, providing proximity to district health administration and specialist services while retaining the resource constraints typical of Ugandan districts.

### 2.3 Quantitative Component

#### 2.3.1 Participants and Sampling

Deaf adolescents aged 13 to 19 years enrolled at WSSD were eligible to participate. Inclusion criteria were: confirmed deafness or severe hearing impairment, age within the specified range, and provision of informed assent (for participants under 18 years) alongside written informed consent from a parent or guardian. Consecutive sampling was employed until the target sample was reached. A target sample size of 68 was calculated based on an estimated SRH knowledge prevalence of 50%, a 95% confidence interval, and a precision of 10%. A final sample of 70 was achieved.

#### 2.3.2 Data Collection Instrument

Data were collected using a structured questionnaire covering: (i) socio-demographic characteristics; (ii) knowledge of SRH components including puberty, contraception, STIs, HIV/AIDS, menstrual health, and gender-based violence; (iii) perceived need for SRH education and services; (iv) sources of SRH information received; and (v) self-reported barriers to accessing SRH services. The instrument was translated into Ugandan Sign Language (UgSL) and administered by a trained deaf research assistant. It was piloted with five deaf adolescents outside the main study site and refined accordingly.

### 2.4 Qualitative Component

#### 2.4.1 Key Informant Selection and Interviews

Four key informants were purposively selected based on their institutional roles and direct engagement with deaf adolescents’ SRH or educational needs: the District Inspector of Schools for Special Needs Education (SNE) in Wakiso District; the Headteacher of WSSD; the School Nurse at WSSD; and the Special Needs Officer for Wakiso District. Semi-structured KIIs were conducted in English using a guide covering: (i) perceived SRH needs; (ii) existing platforms and mechanisms for SRH delivery; and (iii) challenges faced by deaf adolescents in accessing SRH services. Interviews were audio-recorded with permission, transcribed verbatim, and reviewed against original recordings for accuracy.

#### 2.4.2 Focus Group Discussion with Deaf Adolescents

A focus group discussion was conducted with deaf adolescent students at WSSD to centre their first-person experiential perspectives. Participants were purposively selected to include a mix of ages and sexes representative of the student population. The FGD was facilitated by a trained deaf facilitator using Ugandan Sign Language, with a hearing note-taker and interpreter present. The FGD guide covered: (i) understanding of SRH; (ii) knowledge of pubertal changes; (iii) factors motivating SRH service-seeking; (iv) barriers to seeking services; (v) experiences at health facilities or with the school nurse; and (vi) alternative sources of SRH information. The FGD was audio-recorded, transcribed verbatim, and translated into English for analysis.

#### 2.4.3 Qualitative Analysis

All qualitative transcripts (KIIs and FGD) were analysed using thematic analysis following the six-phase framework of Braun and Clarke [6]: familiarisation with data, generating initial codes, searching for themes, reviewing themes, defining and naming themes, and producing the report. Two researchers independently coded the transcripts and met to resolve discrepancies through discussion. Final themes and sub-themes were agreed upon by consensus. Illustrative participant quotes anchor themes throughout the results section.

### 2.5 Integration of Quantitative and Qualitative Data

Integration was conducted at the interpretation phase using a triangulation protocol. Quantitative findings were compared with qualitative themes from both KIIs and the FGD to assess convergence, divergence, and complementarity. Where qualitative data elaborated on or provided mechanisms for quantitative findings, this was explicitly noted. The FGD findings were further compared with KII themes to assess alignment and divergence between institutional and adolescent perspectives.

### 2.6 Ethical Considerations

Ethical approval was obtained from Mildmay Uganda Research Ethics Committee (MUREC), approval number MUREC-2022-96 on 20^th^ April 2020 and the Uganda National Council of Science and Technology, approval number HS2250ES, prior to study commencement. Written informed consent was obtained from parents or guardians of participants below 18 years, and written assent from all participants. Key informants provided written consent for interview participation and recording. FGD participants provided assent and guardian consent as appropriate. Participation was entirely voluntary; participants could withdraw at any time without consequence. Data were anonymised at the point of collection. Deaf research assistants and facilitators were used throughout to ensure linguistic accessibility and cultural safety.

### 2.7 Quantitative Data Analysis

Data were entered into a password-protected electronic database and cleaned prior to analysis. Descriptive statistics were generated for all study variables. Continuous variables are reported as means with standard deviations (SD); categorical variables as frequencies and proportions. All quantitative analyses were conducted using Stata version 15 (StataCorp, TX, USA).

## 3. Results

### 3.1 Quantitative Findings

#### 3.1.1 Socio-Demographic Characteristics

A total of 70 deaf adolescents were enrolled. The mean age was 17.0 years (SD ±1.8), with ages ranging from 13 to 19 years. Females constituted the majority (n=46; 65.7%), with males accounting for 34.3% (n=24). All participants were enrolled at WSSD and communicated primarily through Ugandan Sign Language.

#### 3.1.2 Knowledge of Sexual and Reproductive Health

Of the 70 participants, 88.6% (n=62) reported having heard of at least one SRH component. Components most commonly known included puberty and body changes and menstrual health, while knowledge of contraceptive methods and STI transmission routes was less consistent. Eleven percent (n=8) had not heard of any SRH component.

#### 3.1.3 Perceived Need and Access to SRH Services

An overwhelming majority of 98.6% (n=69) reported a perceived need for SRH education or services. Despite near-universal need, access remained severely constrained: 84.3% (n=59) reported challenges in accessing SRH services at least 85% of the time they sought to do so. Barriers spontaneously cited by participants included absence of sign language interpreters at health facilities, provider discomfort with deaf patients, absence of deaf-appropriate educational materials, transport challenges, and service costs.

#### 3.1.4 Sources of SRH Information

Among participants, 46.0% (n=32) reported having received some form of SRH education. Sources were predominantly informal: friends and peers (45.7%, n=32), parents or guardians (25.7%, n=18), teachers (20.0%, n=14), and the internet or social media (11.4%, n=8). Critically, no participant (0.0%) reported accessing SRH education or services through a government or private health facility. Table 1 presents a full summary of participant characteristics and key findings.

**Table 1.** Participant Characteristics and SRH Knowledge and Access Findings (N=70)

| Variable | Category | n | % | Mean (SD) |
| --- | --- | --- | --- | --- |
| Age (years) | 13–19 | 70 | — | 17.0 (±1.8) |
| Sex | Female | 46 | 65.7% | — |
|  | Male | 24 | 34.3% | — |
| Heard of SRH components | Yes | 62 | 88.6% | — |
|  | No | 8 | 11.4% | — |
| Perceived need for SRH | Yes | 69 | 98.6% | — |
|  | No | 1 | 1.4% | — |
| Challenges with access | Yes (≥85% of the time) | 59 | 84.3% | — |
| Source of SRH information* | Friends/Peers | 32 | 45.7% | — |
|  | Parents/Guardians | 18 | 25.7% | — |
|  | Teachers | 14 | 20.0% | — |
|  | Internet/Social Media | 8 | 11.4% | — |
|  | Government/Private Facility | 0 | 0.0% | — |

*SD = Standard Deviation; SRH = Sexual and Reproductive Health. \*Percentage exceeds 46% due to multiple responses. Percentages for sources calculated as a proportion of total sample (N=70).*

### 3.2 Qualitative Findings: Key Informant Interviews

Five themes emerged from thematic analysis of the four key informant interviews. Table 2 provides an overview of themes, sub-themes, and illustrative quotes.

**Table 2.** Thematic Summary of Key Informant Interview Findings.

| Theme | Sub-themes | Illustrative Quote |
| --- | --- | --- |
| Communication Barriers | Lack of sign language at health facilities; language isolation at home; reliance on hearing siblings as intermediaries | "Students visit hospitals with their parents and no one can understand their communication." — Headteacher |
| Inadequate & Inaccessible Services | Absence of trained SRH staff; negative provider attitudes; no one-on-one or small-group sessions | "Organisations that come do not have time for smaller individual sessions yet this kind of intervention needs a one on one." — School Nurse |
| Family & Community Isolation | Parents locking deaf adolescents at home; fathers struggling to engage; information passed only through hearing siblings | "Deaf adolescents are left behind; many are locked in houses by parents in a bid to protect them." — School Nurse |
| Existing Platforms & Positive Practices | School family initiative; senior women/men teachers; NGO partnerships; reusable pad training; church fellowships | "We have partnered with NGOs who have provided services to deaf adolescents, by training the girl child in making reusable pads." — District Inspector |
| Provider Attitudes & Capacity | Negative attitudes among some health workers; limited skills despite some training; need for openness and befriending approaches | "The health service providers still have negative attitudes though some were trained." — District Inspector |

KII = Key Informant Interview. Quotes are from audio-recorded, verbatim transcripts. Speaker identifiers have been used with participant consent.

#### Theme 1: Communication Barriers as the Primary Structural Obstacle

All four key informants identified communication as the most pervasive barrier to SRH access for deaf adolescents, operating at multiple levels: within the home, in the community, and most acutely within health facilities. The Headteacher described the clinical dimension with particular clarity:

> *“First of all, there is a barrier in language; when the students go back home they face a communication problem because many of their parents and family members cannot use sign language. They have even told us that they look forward to returning to school because here there is live communication. Students express their plight when they visit hospitals with their parents and no one can understand their communication*.*”* — Headteacher, WSSD

The School Nurse confirmed that communication deficits shaped not only access but also the safety and dignity of clinical encounters. The District Inspector of Schools explicitly named the lack of sign language skills among parents and health personnel as a barrier, alongside negative attitudes toward deaf individuals in service settings.

#### Theme 2: Inadequate and Inaccessible SRH Services

Key informants consistently described SRH services as not designed for deaf adolescents— either in format, delivery mode, or provider competency. The School Nurse noted that organisations visiting the school for SRH education typically conducted large-group sessions incompatible with the communication needs of deaf learners:

> *“Some organisations have come but the problem is they do not have time for smaller individual sessions, yet this kind of intervention needs a one on one or smaller groups*.*”* — School Nurse, WSSD

The District Inspector confirmed that health education personnel lacked skilled staff to engage deaf clients, and that negative attitudes persisted even after training. The Special Needs Officer underscored that guidance and counselling services were effectively limited to students enrolled in formal schools, leaving out-of-school deaf adolescents without structured access.

#### Theme 3: Family and Community Isolation

Key informants described how deaf adolescents are frequently isolated from SRH information within their own families. The most common pattern was the routing of information exclusively through hearing siblings:

> *“At home many parents provide this information through the hearing siblings only and they rely on them to tell the deaf*.*”* — School Nurse, WSSD

A more extreme form of isolation—physical confinement within the home by parents seeking to protect deaf adolescents—was also described, paradoxically deepening social exclusion and limiting life skills acquisition:

> *“Deaf adolescents are left behind; many are locked in houses by parents in a bid to protect them, but they need to know and understand how to interact with the society around them and acquire life skills to manage in a hearing world*.*”* — School Nurse, WSSD

#### Theme 4: Existing Platforms and Positive Practices

Key informants identified existing platforms and promising practices that reach deaf adolescents with some SRH education, including a school-based trio of senior women teachers, girls’ matrons, and the school nurse; the School Family Initiative; NGO partnerships delivering practical SRH content; and church fellowship groups. These demonstrate that reaching deaf adolescents with SRH content is feasible when communication is planned for and when trusted community structures are engaged. However, none of these platforms are health facilities, and their coverage and quality remain uneven.

#### Theme 5: Provider Attitudes and Capacity

A recurring concern across interviews was the attitudinal and competency gap among health and education providers. The District Inspector noted that negative attitudes persisted even after training, suggesting that one-off sensitisation efforts are insufficient. The School Nurse framed the necessary shift not only as technical but as relational, a move toward openness and befriending as a professional stance:

> *“From my experience escorting many deaf students and staff to hospitals I have realised that they need openness and friendship; to be friendly to them*.*”* — School Nurse, WSSD

### 3.3 Qualitative Findings: Focus Group Discussion with Deaf Adolescents

The FGD with deaf adolescent students yielded six thematic areas that directly complement and extend the KII findings, while adding the critical dimension of adolescents’ own voices and experiences. Table 3 presents a structured summary of FGD themes, representative responses, and analytical notes.

**Table 3.** Thematic Summary of Focus Group Discussion Findings (Deaf Adolescents)

| FGD Theme | Key Participant Responses | Analytical Note |
| --- | --- | --- |
| Understanding of SRH | Caring about our bodies<br>Protecting ourselves from HIV<br>Minding about our privacy<br>Our personal health | Conceptualisation limited predominantly to body hygiene and HIV prevention; broader dimensions (contraception, GBV, reproductive rights) absent from spontaneous responses. |
| Knowledge of Pubertal Changes | Pubic hair, menstruation, body odour<br>Development of the body<br>Wet dreams<br>Enlargement of breasts and buttocks | Participants could identify physical changes for both sexes but were unable to explain the hormonal or physiological mechanisms underlying them. |
| Motivators for Seeking SRH Services | Pain during menstruation<br>Irregular periods or amenorrhoea<br>Unexplained body odour<br>Confusing bodily feelings<br>Teacher unable to explain; sought school nurse<br>Concern about breast size | Service-seeking was reactive and symptom-driven. No participant described proactive or preventive SRH engagement. The school nurse emerged as the primary accessible touchpoint. |
| Barriers to Seeking SRH Services | Communication barriers (no sign language at facilities) | *"I very much want to seek SRH services but I don't trust people"* |
|  | Shyness and fear of exposure<br>Difficulty expressing feelings in words<br>Lack of confidence<br>Fear of health officers<br>Absence of support persons<br>Mistrust of providers<br>Oppression / difficulty opening up | *Barriers were both structural (communication) and psychosocial (trust, shame, self-efficacy). Several participants described an explicit desire for services alongside an inability to access them.* |
| Experiences at Health Facilities / School Nurse | School nurse provided: body hygiene, emotional self-regulation, guidance and counselling, openness, school regulations<br>No participant had visited Wakiso Health Centre<br>One participant visited a clinic with mother — staff did not know sign language | The school nurse was the sole functional SRH access point. Government health facilities were universally inaccessible due to communication barriers. |
| Alternative Information Sources | Internet; teachers; mothers; friends | Sources mirror quantitative findings. Reliance on informal channels reflects structural absence of formal accessible SRH provision. |

*FGD = Focus Group Discussion. Responses are from verbatim transcription facilitated in Ugandan Sign Language and translated into English. All identifying information has been removed.*

#### FGD Theme 1: Narrow Conceptualisation of SRH

When asked to explain what they understood by SRH, participants consistently centred their responses on body hygiene and HIV prevention. Representative responses included: ‘it means caring about our bodies’, ‘protecting ourselves from HIV’, and ‘minding about our privacy’. While these responses reflect some exposure to health messaging, they reveal that participants’ understanding of SRH does not extend to its broader dimensions: contraception, gender-based violence, reproductive rights, or mental and emotional health aspects of sexuality. This narrow conceptualisation reflects the limited scope of the SRH content participants had been exposed to, primarily through peers, school staff, and internet searches, rather than formal comprehensive SRH education.

#### FGD Theme 2: Partial Knowledge of Pubertal Changes

Participants were able to identify a range of pubertal changes for both sexes, including pubic hair growth, menstruation, body odour, breast and buttock enlargement, and wet dreams. This demonstrates a baseline of experiential awareness. However, participants were unable to explain the hormonal or physiological mechanisms underlying these changes, and could not articulate the relationship between pubertal development and reproductive health or sexual rights. This finding is consistent with the quantitative data showing that knowledge of specific SRH components was uneven, and with the KII observation that existing SRH education at the school is limited in depth.

#### FGD Theme 3: Reactive and Symptom-Driven Service-Seeking

The motivators participants described for seeking SRH services were uniformly reactive and symptom-driven. Reasons included pain during menstruation, irregular periods or amenorrhoea, unexplained bodily feelings, and concerns about breast size or body odour. No participant described proactively seeking preventive SRH information or services. Significantly, the school nurse emerged as the primary and often sole accessible touchpoint. One participant described escalating to the nurse after a teacher was unable to answer their question, illustrating how the school-based care pathway functions as a sequential last resort rather than a first point of contact.

#### FGD Theme 4: Multilevel Barriers to Service-Seeking

Participants identified a complex constellation of barriers to seeking SRH services, spanning structural and psychosocial dimensions. Communication barriers were foremost: ‘people do not know sign language’ was explicitly named, and the inability to express bodily feelings in words, particularly technical or emotional terms, was described as paralyzing. Participants also described fear of health officers, absence of trusted support persons, shyness, and lack of self-confidence. Most strikingly, one participant articulated the paradox of simultaneous desire and inability:

> *“I very much want to seek SRH services but I don’t trust people*.*”*

This statement captures a recurring dynamic across FGD responses: access is not primarily blocked by lack of motivation or awareness, but by the intersection of structural inaccessibility with psychosocial vulnerability. Several participants described a sense of oppression that made it difficult to open up, and others described the impossibility of explaining their experiences when they lacked the vocabulary to do so in any language accessible to hearing providers.

#### FGD Theme 5: The School Nurse as the Sole Functional Access Point

Participants who had sought SRH-related help reported that the school nurse provided information on body hygiene, emotional self-regulation, guidance and counselling, openness, and school regulations. No participant had ever visited Wakiso Health Centre for SRH concerns. One participant reported attending a clinic with her mother but was not served because staff did not know sign language. These findings confirm the quantitative finding that no participant had accessed SRH through a formal health facility, and provide qualitative explanation: the government health facility is structurally inaccessible, while the school nurse, despite limited capacity and scope, functions as the de facto provider of SRH support for this population.

#### FGD Theme 6: Reliance on Informal and Alternative Information Sources

In the absence of formal SRH services, participants reported seeking information from the internet, teachers, mothers, and friends. This mirrors the quantitative finding that peers and informal networks are the dominant information sources. The internet was notable as an autonomous, anonymous channel; however, its utility is constrained by variable literacy, limited data access, and the absence of content in Ugandan Sign Language. Reliance on friends, while common, introduces risks of misinformation and peer pressure that structured peer-education programmes could help mitigate.

### 3.4 Integration: Convergence and Complementarity Across Three Data Strands

The quantitative, KII, and FGD findings are highly convergent and mutually reinforcing, while each adding distinct value. The quantitative finding that no participant accessed SRH through a formal health facility is directly confirmed and explained by both the KII themes of communication barriers and provider attitudes, and the FGD accounts of inaccessible government facilities and the singular role of the school nurse. The quantitative reliance on informal sources is elaborated qualitatively through the FGD descriptions of internet use, peer consultation, and maternal engagement, and contextualised by the KII themes of family isolation and existing school-based platforms.

The FGD adds an essential layer absent from KII data alone: the psychosocial texture of access barriers from adolescents’ own perspectives. The emotional dimensions of shyness, mistrust, oppression, and the inability to articulate bodily experience, captured in FGD data, explain why structural reforms alone (such as provision of sign language interpreters) may be insufficient without simultaneous attention to adolescents’ confidence, trust, and communication self-efficacy. The FGD also reveals that the narrow scope of adolescents’ SRH conceptualisation is not a product of disinterest, but of limited and shallow exposure to SRH content.

Together, the three data strands confirm that the SRH access gap for deaf adolescents in Uganda is structural, multilevel, and psychosocially embedded. It is unlikely to close without deliberate action across health systems, families, schools, and community environments.

## 4. Discussion

This mixed-methods study provides among the first empirical evidence on SRH knowledge and access for deaf adolescents in Uganda, combining quantitative prevalence data with rich institutional perspectives from key stakeholders and, critically, the first-person voices of deaf adolescents through an FGD. Our findings reveal a well-recognised, in disability and health equity literature: high levels of perceived need coexist with profound structural barriers to accessing appropriate services [8]. That 98.6% of participants felt a need for SRH education or services and yet 84.3% could not access them in the majority of their attempts, and also that none had ever received SRH from a health facility, points to a systemic failure rather than a deficit of motivation or interest among deaf adolescents themselves.

The FGD finding that participants’ conceptualisation of SRH was narrow and limited to body hygiene and HIV prevention, reflects the underprivileged scope of their SRH exposure rather than cognitive limitation. International evidence consistently demonstrates that when deaf adolescents are provided with accessible, comprehensive, and appropriately formatted SRH education, their knowledge and protective behaviours improve significantly [9]. The current study documents the conditions that prevent such exposure from occurring in Uganda.

The complete absence of health facilities as an SRH information source is the most striking quantitative finding, and it receives contextual explanation from both the KIIs and the FGD. Key informants across four institutional roles independently described health facilities as functionally inaccessible to deaf adolescents: lacking sign language interpreters, staffed by providers with negative or fearful attitudes, and not designed for individual or small-group communication. FGD participants confirmed this from their own experience, with one participant recounting being unserved at a clinic due to the absence of sign language communication. The FGD data further reveal that beyond structural inaccessibility, psychosocial barriers including mistrust of providers, fear, shame, and the inability to articulate bodily experience, compound the structural exclusion.

The communication barrier identified as primary across KII and FGD data has both technical and attitude dimensions. Technically, the absence of qualified sign language interpreters renders the clinical encounter inaccessible. Aon the attitude aspect, key informants noted that negative attitudes toward deaf patients persisted even after training, pointing to the insufficiency of one-off sensitisation efforts. The School Nurse’s framing of ‘openness and friendship’ as the necessary relational posture offers a useful, person-centred principle for health worker training. The FGD amplifies this: participants articulated that the absence of trust and the presence of fear of health officers were decisive deterrents to seeking care.

Family and community isolation constitutes a second major driver of the access gap. The routing of SRH information through hearing siblings is an inherently unreliable and dignity-compromising system. The FGD confirms that deaf adolescents are aware of their own need for direct communication and direct access to information. They are not passive recipients of a relay system, but active seekers of knowledge who are structurally blocked from accessing it directly. This family-level failure reinforces the quantitative finding that peers constitute the most common source of SRH information, a pattern that carries risks of misinformation alongside potential for structured peer-led education.

The predominantly female composition of our sample (65.7%) mirrors enrolment patterns at the study school and reflects the compounded vulnerabilities at the intersection of gender inequality and disability-related exclusion [10]. The FGD finding that menstrual concerns including pain, irregularity and amenorrhoea were the primary drivers of SRH service-seeking underlines the specifically gendered nature of deaf adolescents’ SRH needs and the inadequacy of a service environment that cannot address even these basic concerns accessibly.

The symptom-driven nature of service-seeking documented in the FGD is a finding with significant programmatic implications. It suggests that deaf adolescents lack access to the proactive, preventive framing of SRH that comprehensive sexuality education provides. The school nurse, their de facto primary provider, is equipped to address immediate concerns but is not positioned to deliver comprehensive SRH education. Linking school-based care to a referral pathway that includes community health workers trained in sign language could extend both the scope and continuity of SRH support.

Several study limitations should be acknowledged. The cross-sectional design precludes causal inference. Data were collected from a single school in one district, limiting generalisability to deaf adolescents in rural areas or those not enrolled in formal education. Self-reported data are subject to social desirability bias on sensitive SRH topics. The FGD captured a subset of students; perspectives may not be fully representative of the broader deaf adolescent population. Future research should employ longitudinal and participatory designs, include diverse geographic settings and out-of-school populations, use validated knowledge instruments, and investigate the feasibility and effectiveness of specific SRH intervention models co-designed with deaf adolescents.

## 5. Conclusion and Recommendations

Deaf adolescents in Wakiso District, Uganda, demonstrate high awareness of SRH and near-universal perceived need for SRH education and services. Yet structural barriers, rooted in communication inaccessibility, inadequate provider training, family isolation, and exclusion from formal health systems, interact with psychosocial barriers of mistrust, shame, and communicative self-doubt to produce profound and multilevel access gaps. No deaf adolescent in this study received SRH services from a health facility. The school nurse carries an inequitable and insufficient burden as the de facto primary SRH provider, while informal networks of peers, parents, and teachers serve as surrogate and inadequate SRH educators.

The integration of quantitative, KII, and FGD findings produces a coherent and actionable evidence picture. On the basis of these findings, we make the following recommendations:

- The Uganda Ministry of Health should mandate disability-inclusive SRH within the national essential health care package, with specific provisions for trained sign language interpretation at all district-level health facilities.
- Health worker pre-service and in-service training curricula should include sustained, competency-based modules on accessible communication with deaf patients, going beyond one-off sensitisation to address attitudinal transformation.
- National SRH education materials should be developed in Ugandan Sign Language and made freely available across schools, communities, and digital platforms.
- Community-level SRH education should be extended to parents, guardians, and siblings of deaf adolescents, with particular attention to fathers and to peri-urban and rural communities where communication isolation is greatest.
- The School Family Initiative and existing school-based platforms should be formally linked to referral pathways at local health facilities to create a continuum of care.
- Future SRH programmes should be co-designed with deaf adolescents, using participatory approaches that centre their expertise, lived experience, and psychosocial needs alongside structural reform.
- Psychosocial dimensions of SRH access, including trust-building, emotional safety, and communication self-efficacy should be explicitly addressed in deaf adolescent SRH programming, not treated as secondary to structural change.

Deaf adolescents must be recognised as rights-holders with full entitlement to comprehensive, accessible, and dignified SRH care, not as an afterthought, but as a central priority in Uganda’s health equity agenda [7].

## Data Availability

All quantitative data are contained within the manuscript and its supporting information. Qualitative data (interview transcripts and focus group discussion transcripts) cannot be shared publicly due to the sensitivity of the topic and to protect participant confidentiality, as participants did not consent to public data sharing. Anonymised qualitative data may be made available to qualified researchers upon reasonable request to the corresponding author at.

## Acknowledgements

The authors acknowledge the National Institute for Health Research (NIHR) and the Royal Society of Tropical Medicine and Hygiene (RSTMH) for funding this study through the RSTMH Small Grants 2021 Programme. The views expressed are those of the authors and not necessarily those of the NIHR or RSTMH.

We are grateful to the district officials of Wakiso District, including the District Inspector of Schools for Special Needs Education and the Special Needs Officer, for their cooperation and facilitation of this research. We thank the administration and staff of Wakiso Secondary School for the Deaf for granting access and providing institutional support throughout the study period.

We extend our deepest appreciation to the deaf adolescent participants who gave their time, trust, and voices to this study. Their willingness to share their experiences is the foundation of this work. We also thank the key informants—the Headteacher, School Nurse, District Inspector of Schools, and Special Needs Officer—for their candid insights.

We acknowledge the invaluable contribution of the research assistants, particularly the deaf research assistant who administered the quantitative questionnaire in Ugandan Sign Language and the deaf facilitator who led the focus group discussion. Their linguistic expertise and community trust were essential to the integrity of data collection.

## References

1. World Health Organization. World Report on Disability. Geneva: WHO; 2011.

2. United Nations. Transforming Our World: The 2030 Agenda for Sustainable Development. New York: UN; 2015. Resolution A/RES/70/1.

3. Banks LM, Davey C, Shakespeare T, Kuper H. Disability-inclusive responses to COVID-19: Lessons learnt from research and practice. World Dev. 2021;137:105178.

4. Uganda Bureau of Statistics. Uganda National Population and Housing Census. Kampala: UBOS; 2014.

5. Uganda Ministry of Health. National Adolescent Health Policy. Kampala: MoH; 2020.

6. Braun V, Clarke V. Using thematic analysis in psychology. Qual Res Psychol. 2006;3(2):77–101.

7. Shakespeare T. Disability Rights and Wrongs Revisited. 2nd ed. London: Routledge; 2014.

8. Groce N, Challenger E, Berman-Bieler R, et al. Malnutrition and disability: unexplored opportunities for collaboration. Paediatr Int Child Health. 2014;34(4):308–314.

9. Kuper H, Heydt P. The Mission Billion: Access to Health Services for 1 Billion People with Disabilities [Internet]. London: London School of Hygiene and Tropical Medicine; 2019 [cited 2024 Jan]. Available from: https://www.lshtm.ac.uk/research/centres/international-centre-evidence-in-disability/news/102251/mission-billion

10. World Health Organization. Women and Health: Today’s Evidence, Tomorrow’s Agenda. Geneva: WHO; 2009.

